# The causal relationship between cathepsins and multiple sclerosis : a mendelian randomization study

**DOI:** 10.1101/2024.09.05.24313125

**Authors:** Haining Lin, Yuqing Shi, Huazhong Xiong, Dongyi Wang, Shilei Wang, Hailan Kang, ZiXu Wang, Zeyu Wang, Jixiang Ren

## Abstract

**Background:** Multiple sclerosis (MS) is a chronic autoimmune neurodegenerative disease. Although some evidence indicates the potential involvement of cathepsins in the MS process, the precise nature of this involvement remains uncertain. The objective of this study was to ascertain whether there is a causal relationship between cathepsins and MS.

**Methods:** This study aimed to examine the relationship between nine cathepsins and MS, incorporating heterogeneity and sensitivity analyses into the study design. The MR study was reported according to the STROBE-MR checklist.

**Results:** The MR analysis revealed a causal relationship between cathepsin H and MS (IVW: P=0.036, OR=1.095, 95% CI=1.006-1.192); and an inverse causal relationship between cathepsin E and MS (IVW: P=0.031, OR=1.043, 95% CI=1.004-1.083).

**Conclusion:** Genetically predicted risk of developing MS was associated with increased cathepsin H, whereas elevated cathepsin E was associated with developing MS, and their causal relationships were both unidirectional.

## Introduction

Multiple sclerosis (MS) is a chronic inflammatory illness caused by the immune system, characterized by neurodegeneration and demyelination[1].MS has the potential to result in a reduction in the quality of life of those affected, with the possibility of further complications such as disability. This is due to the fact that MS can manifest itself in neurological symptoms.The incidence and prevalence of MS are increasing annually[2].In addition to several well-defined environmental factors. Furthermore, numerous genes have been identified that, to a moderate extent, increase disease susceptibility.[3].Autoimmune activity against myelin components is thought to be the primary process underlying the pathology of MS[4],including significant infiltration of activated leukocytes into the central nervous system(CNS) via the blood-brain barrier and processing and presentation of self-antigens by antigen-presenting cells.[4, 5].

An imbalance between protein hydrolysis and protein synthesis is a key factor in the development of CNS diseases. Consequently, the study of protein degradation pathways is of great importance in understanding the underlying pathology of these diseases[6].Cathepsins are involved in a number of cellular processes, including cell signalling, autophagy, and the transformation of proteins and lipids. They are primarily responsible for the metabolic degradation of peptides and proteins within lysosomes[7, 8].Some evidence suggests that in patients with MS, enzymatic activities of the proteasome are enhanced in the central nervous system.[9].Cathepsins can be classified into three categories: cysteine cathepsins (cathepsin B, C, L, etc.), serine cathepsins (cathepsin G), and aspartate cathepsins (cathepsin D and E)[10]. Furthermore, there is a strong correlation between the activity of tissue proteases and the progression of neurodegenerative diseases[11, 12].In immune cells, cathepsin S and cathepsin L play a pivotal role in the processing of antigenic peptides and the presentation of antigens by histocompatibility complex class II molecules. They are also involved in the progressive catabolism of integral membrane proteins, which are encoded by invariant chains and are expressed on the surface of antigen-presenting cells[13–15].The invasive and migratory properties of certain cathepsins [16, 17]may be attributed to their capacity to maintain activity and degrade extracellular matrix (ECM) components, including laminin, and collagen, at physiological pH values[18].Given that protein hydrolytic degradation constitutes a pivotal process in demyelination during MS, with the potential to precipitate neurodegeneration, cathepsins have been demonstrated to cleave myelin basic proteins, thereby exerting a pivotal role in disease progression[19].

In previous studies, although there is some evidence that cathepsins are involved in the process of MS, there is a lack of clear causality between cathepsins and MS, we used.Mendelian randomisation (MR) analysis, which combines data from genome-wide association studies (GWAS), is a statistical method used to determine the causal relationship between exposures and outcomes[20].MR analyses have been used more widely to assess possible causality between exposures and outcomes because of the effectiveness of MR in reducing confounding bias and preventing reverse causation, advantages that are based on the inability of the disease to alter the genotypes in the germ line.

The goal of this study was to investigate the relationship between nine cathepsins and MS, as identified through a large-scale genome-wide association study (GWAS), through a comprehensive analysis of the genetic variants associated with them. This was accomplished through the utilisation of a two-sample bidirectional MR approach, which yielded insightful findings that significantly advanced the comprehension of the underlying causal relationship between cathepsins and MS.

## Materials and methods

### Data sources

The gene detection levels of cathepsin B, E, F, G, H, L2, O, S, and Z (a total of nine types of cathepsins) were obtained from the INTERVAL study. The data can be accessed publicly via the following website: https://gwas.mrcieu.ac.uk. The sample size of this study is 3,301, and the participants are of European ethnicity[21].The INTERVAL study has been approved by the National Research Ethics Service in the United States, with approval number 11/EE/0538.

The International Classification of Diseases, Eleventh Revision code (ICD-11) for MS is 8A40. The disease is defined as an inflammatory demyelinating disease characterised by episodic neurological dysfunction in at least two central nervous system regions (brain, spinal cord, and optic nerves) at different times and in different spaces (different sites of lesions). The diagnostic criteria follow the 2017 version of the McDonald diagnostic criteria [22]. The GWAS data for MS were obtained from the publicly available GWAS database (https://gwas.mrcieu.ac.uk/), GWAS number ieu-b-18, which analysed the genetic data of 47,429 individuals. A total of 68,374 control subjects and 1,000 individuals with MS were included in the study, which aimed to create a reference atlas of the genetic structure of MS in the European population. The study involved both male and female subjects, and the number of single-nucleotide polymorphisms (SNPs) identified was 6,304,359 [23].

To prevent the introduction of bias and avoid data overlap, the sample data for cathepsins and MS were obtained from independent GWAS databases. No additional ethical approval or licence is required for the study, as all studies undergo review and approval by the relevant institutional ethics review committee and informed written consent is obtained from each participant.

### Study design and selection of IVs

The study methodology was in accordance with the STROBE-MR list [24], and we conducted a two-sample bidirectional MR study using publicly available GWAS summary data.The MR analyses were based on three key assumptions [25], the details of which are shown in Fig. 1, and we performed MR analyses and reverse MR analyses of cathepsins and MS, respectively.

**Fig 1.**
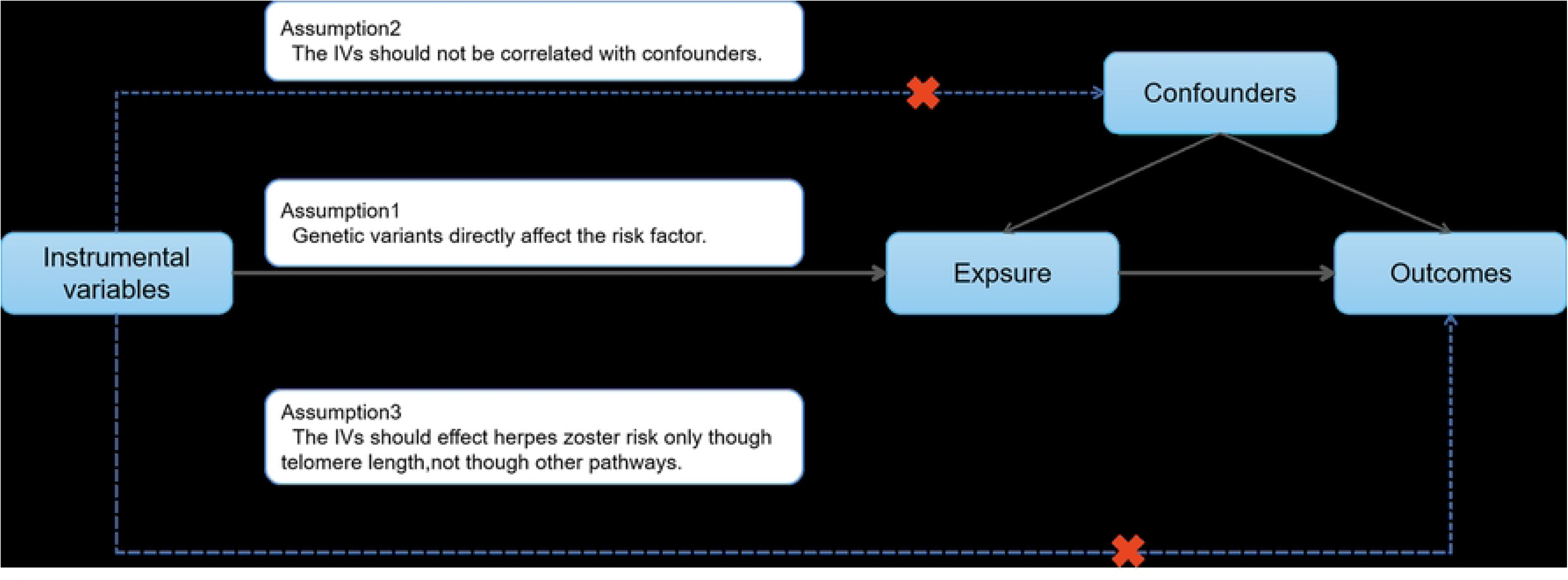
Flowchart of MR analysis. Three key assumptions of MR analysis: (1) IV must be strictly related to the exposure factor; (2) IV must be independent of confounders between the exposure factor and the outcome; and (3) IV will not influence the outcome due to factors other than the exposure factor.

The following methods were employed to identify single-nucleotide polymorphisms (SNPs) associated with exposure factors, with the objective of ensuring the reliability and accuracy of the results. SNPs with robust associations (p-value significance threshold of less than 5 × 10⁻⁶) were treated as IVs. Additionally, those with strand-balanced SNPs (r2 = 0.001, genetic distance = 1000 kb) were excluded to eliminate any potential bond disequilibrium[24]. To evaluate the potential for weak instrumental bias, the F-statistic was employed to assess the weak IV bias of the selected IVs[26].

### MR analysis

The study was analysed using the TwoSampleMR (version 0.5.6) package in R (version 4.4.1), and MR analyses were performed to estimate the overall effect of the selected SNPs on MS. The following MR methods were employed: Egger, Weighted median, Inverse variance weighted, Simple mode and Weighted mode, with the objective of verifying the consistency of the results[27]. Subsequently, inverse MR analysis was conducted using the aforementioned MR methods. The odd ratio (OR) was found to be statistically significant at the 95% confidence level with a two-sided significance test, with P < 0.05.

The MR Egger intercept was employed for the purpose of detecting horizontal pleiotropy. In the event that the intercept was found to be statistically significant (P < 0.05), it was necessary to consider the reliability of the results. The Cochran’s Q statistic was utilised for the assessment of heterogeneity (P < 0.05), while funnel plots were employed for the purpose of The results are illustrated. The MR-PRESSO outlier test and the estimated calibration values were applied to remove horizontal pleiotropy, which can be used to exclude aberrant SNPs. The generation of forest plots in the study enabled a visual assessment of the robustness of the results after SNP removal by the leave-one-out method.

## Results

### IV selection

The present investigation utilised genetic variants from the INTERVAL study to examine cathepsins for MS from the GWAS database. Our bidirectional MR analyses identified multiple SNPs as IVs for each cathepsin and MS disease status, with a genome-wide significance threshold of P<5×10^-6^. The F-statistic for each selected IV exceeded 10, indicating no weak IV bias.

### Results of the main MR analyses

A two-sample MR analysis was conducted to evaluate the impact of cathepsins on MS, employing a range of data analysis techniques, including MR Egger, weighted median, inverse variance weighted, simple mode, and weighted mode. Of these approaches, the inverse variance weighted method is regarded as the most reliable for data analysis, particularly in instances where directed pleiotropy is absent. This method is frequently employed in meta-analyses of the effects of multiple genetic variants.

The findings of the two-sample MR analysis examining the relationship between cathepsins and MS are illustrated in Figure 2. Additionally, a reverse MR analysis investigating the reverse association between MS and cathepsins is presented in Figure 3. All positive results (P > 0.05) are presented in Table 1 for the reader’s convenience. The analysis indicates a causal relationship between cathepsin H and MS, as well as a reverse causal relationship between cathepsin E and MS. Specifically, elevated cathepsin H may be associated with an increased risk of developing MS (IVW: P=0.036, OR=1.095, 95% CI=1.006-1.19). Furthermore, elevated cathepsin E may be associated with the development of MS (IVW: P=0.031, OR=1.043, 95% CI=1.004-1.083).

**Fig 2.**
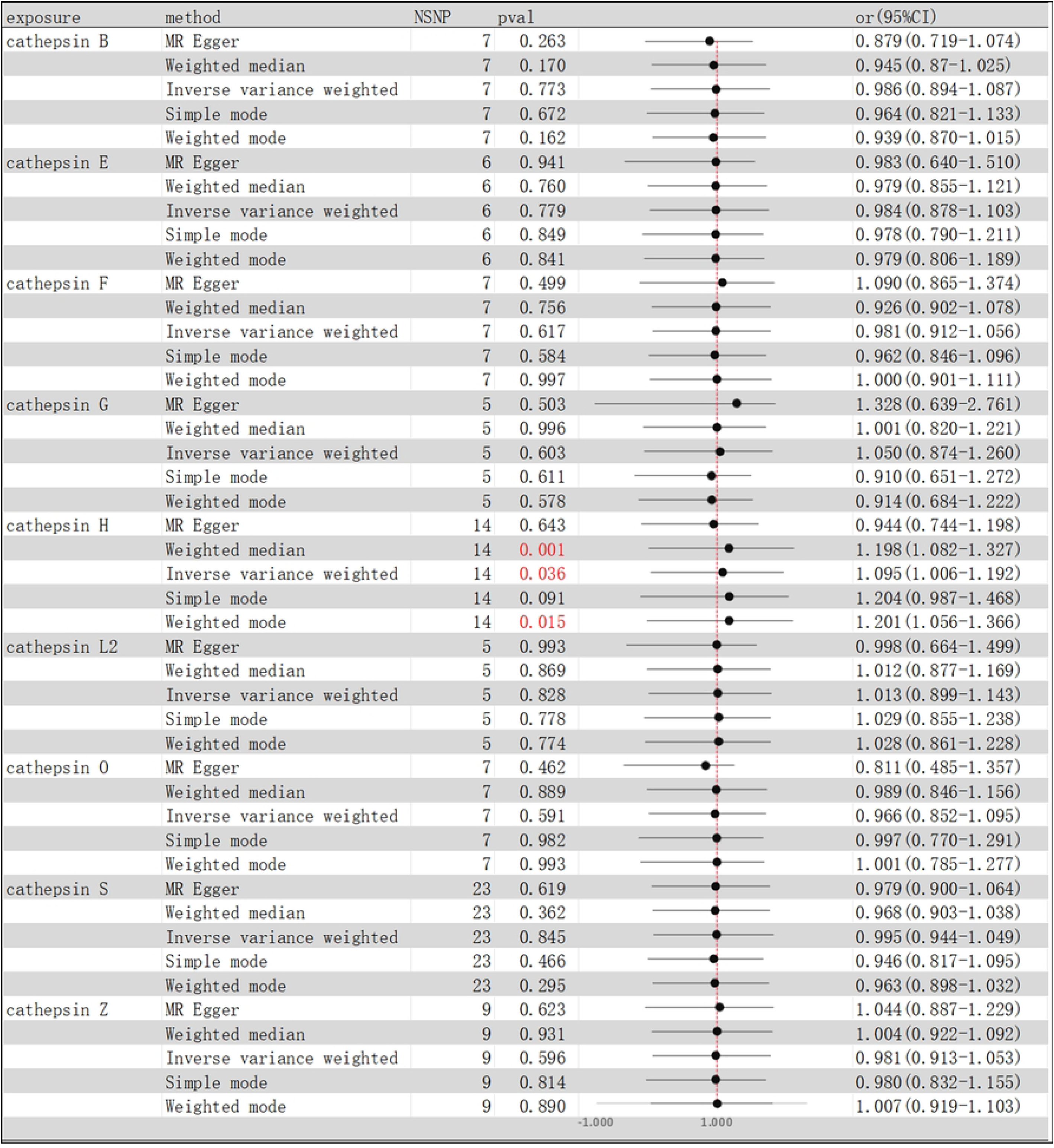
**MR results of the causal relationship between cathepsins and MS,** OR, odds ratio; CI, confidence interval.

**Fig 3.**
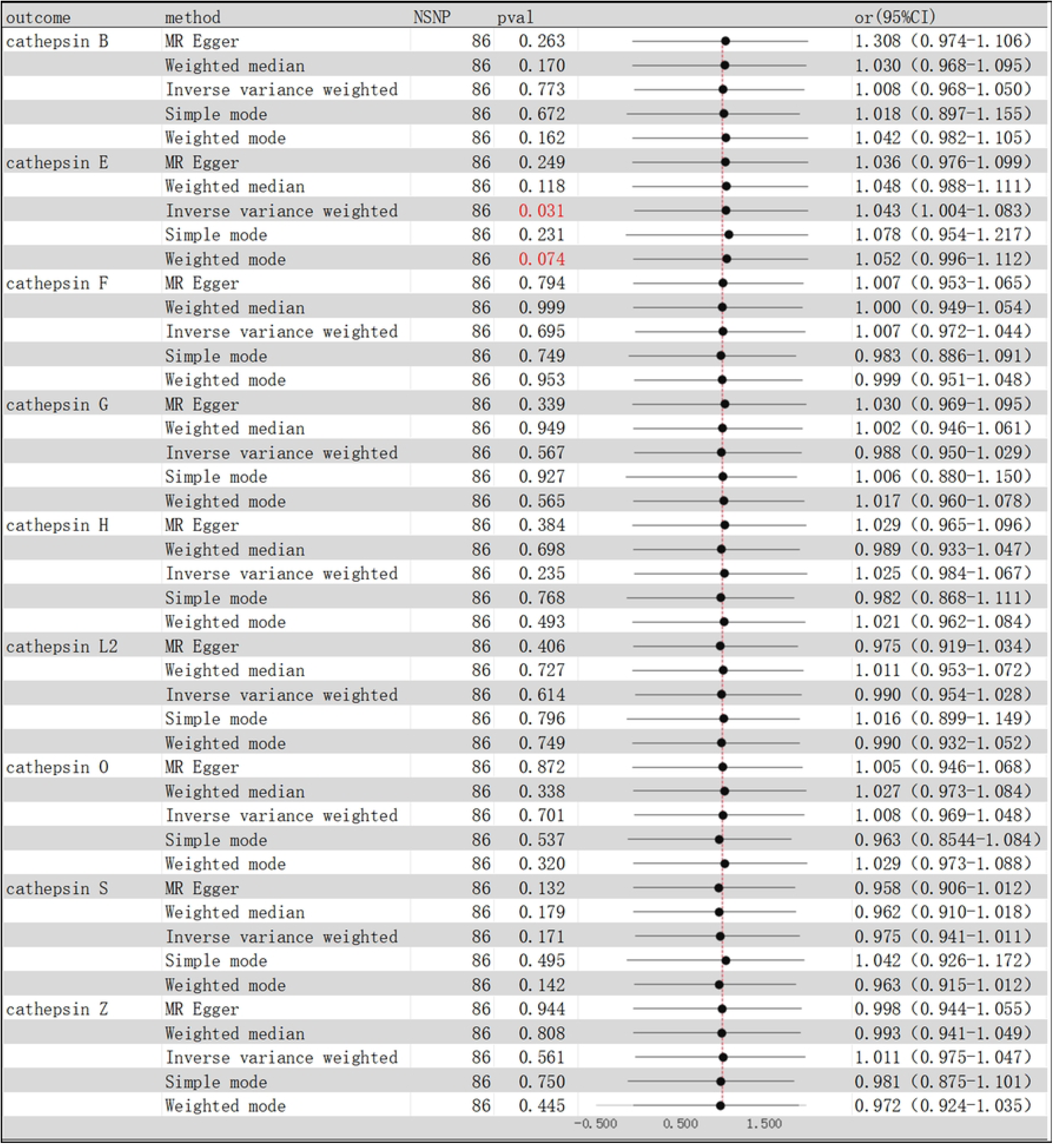
**MR results of inverse causality between cathepsins and MS,** OR, odds ratio; CI, confidence interval.

**Table 1.**
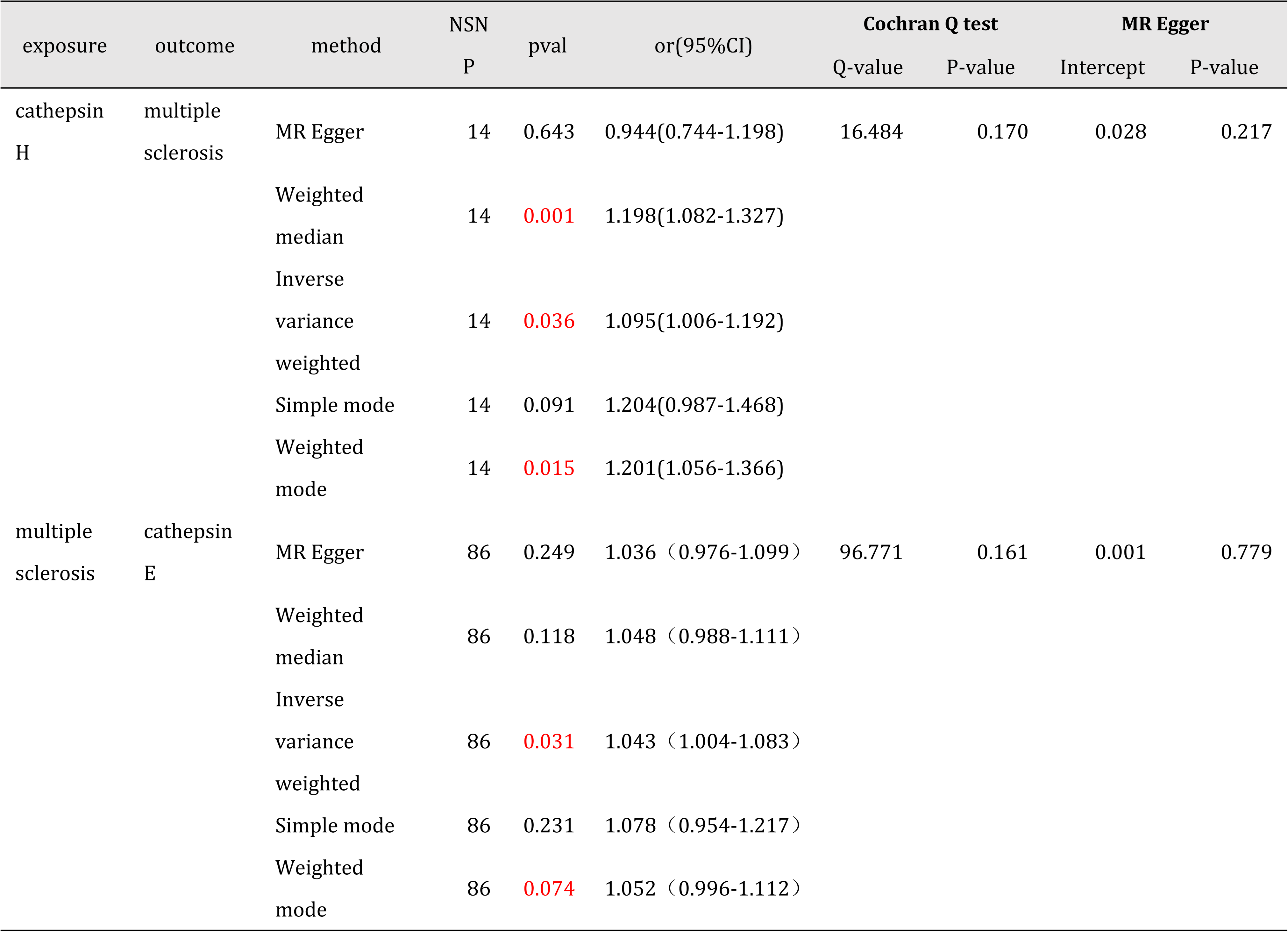
Mendelian randomisation analysis with positive results.

Furthermore, the Cochran Q test was employed to ascertain the extent of heterogeneity between the SNPs. No evidence of heterogeneity or pleiotropy in positive causality was identified(P>0.05), as evidenced by the resulting P-values and MR-Egger intercepts (Table 1). The leave-one-out method analysis additionally validated the robustness of the positive results, as illustrated in Figure 4.

**Fig 4.**
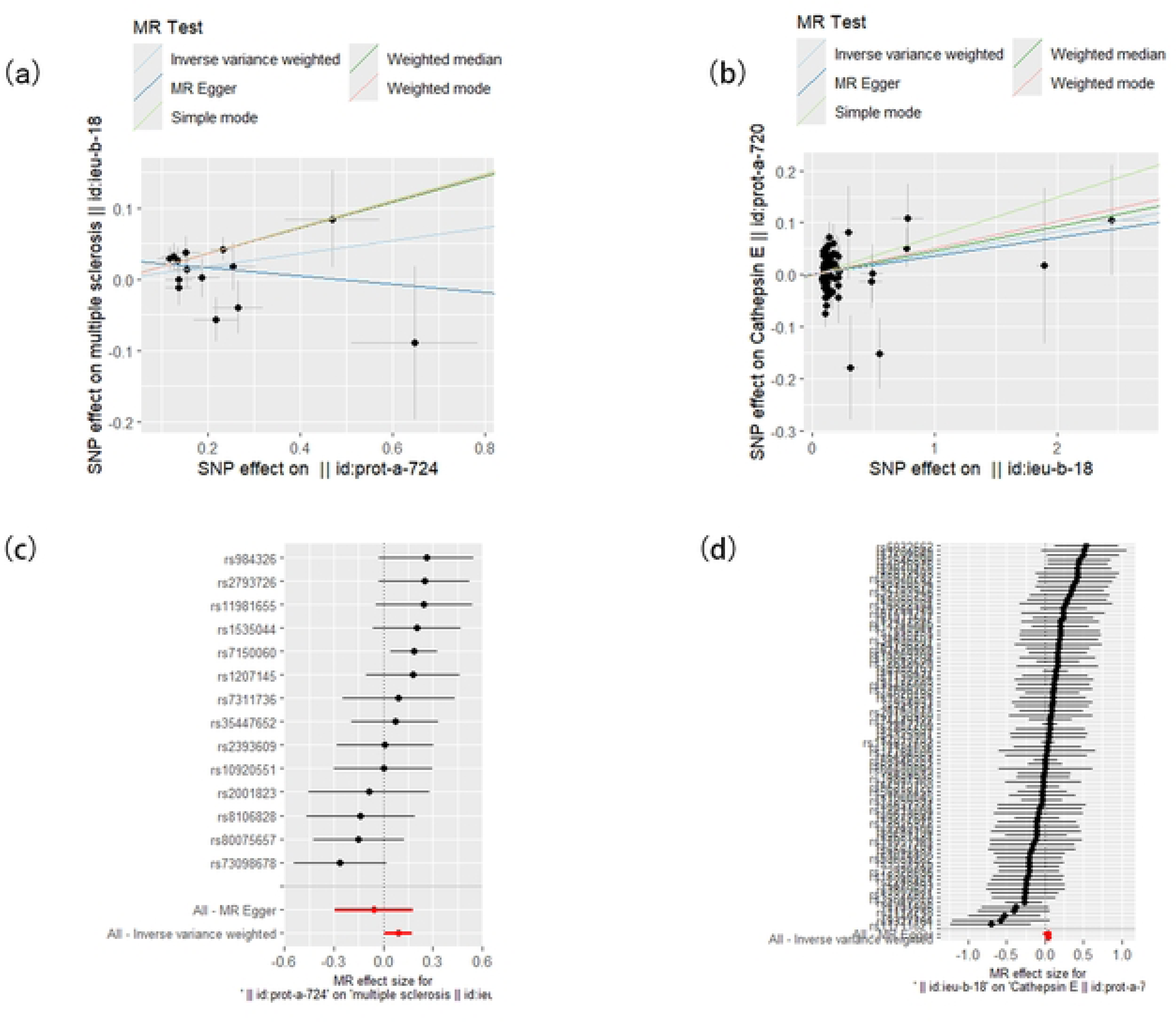
**Scatterplot and leave-one-out analysis of Mendelian randomised causality,** (a) MR analysis of cathepsin H versus MS (b) reverse MR analysis of cathepsin E versus MS (c) sensitivity analysis of the leave-one-out method for cathepsin H versus MS (d) sensitivity analysis of the leave-one-out method for reverse causality of cathepsin E versus MS.

## Discussion

MS manifests itself as an autoimmune demyelinating and neurodegenerative disease of the central nervous system.The mechanism by which MS occurs has not been fully elucidated, and is widely believed to be due to an autoimmune attack on myelin.Genetic and environmental factors are thought to play a role in triggering the immune system’s attack on myelin[28].Cathepsins are associated with the development of neurodegenerative and immunocompetent disorders and are usually correlated with pathological conditions [8, 29]. Evidence from relevant studies has shown [30–33]that a variety of cysteine cathepsins are involved in the inflammatory process of different diseases through different pathological mechanisms [34]. While in autoimmune mechanisms, cathepsins play a major role in phagocytosis and autophagy [35], cathepsins may exert an indirect regulatory effect on autophagy and a direct influence on cellular homeostasis and inflammatory processes[36].In addition, cathepsins play an immunological role in the cytoplasmic lysosol, nucleus or extracellular space, e.g. The release of cathepsins from the cytoplasmic lysosome and nucleus may occur via either controlled or uncontrolled lysosomal membrane permeabilisation, resulting in lysosomal-dependent cell death [37]Conversely, those present in the extracellular space may be involved in the remodelling of the extracellular matrix[10]. Thus cathepsins may be involved in the disease progression of MS.

Our findings suggest that the pathogenesis of MS may be associated with elevated cathepsin H. Although there are fewer studies directly correlating cathepsin H with the pathogenesis of MS, the current mechanism in animal models is thought to be that defective cathepsin H impairs TLR3-mediated activation of IRF3 and the consequent secretion of IFN-β by dendritic cells, leading to enhanced differentiation of Th1 cells, which leads to the early onset of experimental autoimmune encephalomyelitis (EAE, an animal model of multiple sclerosis)[38].However, demyelination is a characteristic manifestation of MS, and myelin is essential for neuronal axonal function in the CNS, whereas microglia regulate the growth and integrity of myelin in the CNS, and are required to maintain myelin integrity by preventing myelin degeneration [39], and microglia are immune cells residing in the brain that are involved in tissue maintenance and immunosurveillance[40]. Cathepsin H was detected in different brain regions within activated microglia, and a related study showed that CatH was significantly upregulated in microglia from mice injected intraperitoneally with lipopolysaccharide, and that pro-inflammatory cytokines induced the release of CatH from microglia, with profound and deleterious effects on microglia activation and neuronal survival[41].Another study showed that cathepsin H expression was increased in microglia at the site of injury in a hypoxic-ischemic mouse model, and that cathepsin H inhibited astrocyte proliferation by controlling TLR3/IRF3 signalling and subsequent IFN-β secretion [42]. We therefore hypothesise that microglia function may be a relevant mechanism for the causal relationship between cathepsin H and the development of MS, however existing studies on the correlation between cathepsin H and MS are insufficient.Further studies are required to gain a comprehensive understanding of the relationship between the aforementioned variables. The findings of the present MR analysis may serve as a valuable point of reference for future investigations into the role of cathepsin H in MS.

In our findings, suffering from MS may lead to elevated cathepsin E. Although evidence that MS directly leads to elevated cathepsin E has not emerged from current research advances, it is interesting to note that related studies have suggested that cathepsin E may be associated with an underlying mechanism of MS-related pain [43]. This study found that cathepsin E-deficient mice were resistant to mechanically abnormal pain in EAE and that cathepsin E contributes to mechanically abnormal pain in EAE mice, however, this study could not confirm the relationship between cathepsin E and MS-related neuropathic pain, as EAE can be induced by a variety of immunogenic peptides, and therefore further analyses are still required. Notably, in the CNS, cathepsin E is expressed only in microglia[44], and microglia activation is a key factor in the induction of mechanical abnormalities in pain[45]; Moreover, cathepsin E has been demonstrated to be a crucial mediator in major histocompatibility complex class II-mediated antigen presentation in microglia [46]. In conclusion, analysis of MR results suggests that suffering from MS may lead to elevated cathepsin E, and related studies suggest that this may be associated with pain in the disease; however, there are fewer available findings on cathepsin E and MS, so further in-depth studies are necessary, and our results may provide potential research directions for future studies on MS-related pain and cathepsin E.

Our findings establish a causal relationship between a range of cathepsins and MS. By employing MR analysis, the data can be understood with greater reliability and accuracy, avoiding potential bias introduced by external factors. MR analysis is a method of analysing genetic variation. The results of the present study contribute to the discovery of the role of cathepsins in the progression of MS and provide further research with potential value. It is important to acknowledge the limitations of this study. Firstly, it should be noted that the GWAS data used in this study were derived from Europe only and therefore do not represent a generalisable sample of the wider population. Consequently, further expansion of the study is still needed. Secondly, there is evidence to suggest that cathepsin B may be associated with MS. A clinical observational study demonstrated an increase in cathepsin B activity in peripheral blood mononuclear cells of MS patients[47], while the use of cathepsin B inhibitors resulted in a significant reduction in the severity of clinical scores in EAE mice [48].However, despite these findings, we were unable to identify a genetic tool for the cathepsin B correlation, which led to the exclusion of our results from the tissue causal analyses of cathepsin B and MS. Secondly, as MR analysis can only provide evidence of potential causality and is unable to identify the specific biological pathways involved, future studies should combine bioinformatics analyses with relevant experiments to elucidate and validate the underlying mechanisms.

## Conclusion

This study examined scathepsins and MS using a bidirectional MR approach and the findings provide suggestive evidence that genetically predicted risk of developing MS is associated with increased cathepsin H and elevated cathepsin E is associated with developing MS, and that they are both unidirectionally causally related, however, further intervention trials are needed to elucidate the underlying mechanisms.

## Data Availability

All relevant data are within the manuscript and its Supporting Information files.

https://gwas.mrcieu.ac.uk/

## Data availability statement

The original contributions from this study are included in the article. Additional data inquiries can be directed to the corresponding authors.

## Author contributions

Haining Lin: Investigation, Data curation, Writing – original draft, Writing – review & editing. Yuqing Shi: Investigation, Data curation, Methodology. Huazhong Xiong: Investigation, Data curation. Dongyi Wang: Investigation, Project administration. Shilei Wang: Investigation, Validation. Hailan Kang: Investigation. Zixu Wang: Investigation. Zeyu Wang:Writing – review & editing.Jixiang Ren: Conceptualization, Writing – review & editing.

## Acknowledgments

The authors are thankful to all participants and investigators who contributed and shared summary-level data on GWAS.

